# AI-literacy training enhances physician-LLM diagnostic collaboration in a resource-limited setting: a randomized controlled trial

**DOI:** 10.1101/2025.06.06.25329104

**Authors:** Ihsan Ayyub Qazi, Ayesha Ali, Asad Ullah Khawaja, Muhammad Junaid Akhtar, Ali Zafar Sheikh, Muhammad Hamad Alizai

## Abstract

Diagnostic errors remain a pervasive yet preventable source of patient harm, with resourcelimited healthcare systems in low- and middle-income countries (LMICs) facing disproportionately higher diagnostic error rates due to limited access to diagnostic tools, specialists, and decision support systems. Large language models (LLMs) offer potential to bridge diagnostic gaps in these settings but can generate inaccurate information, making comprehensive AI-literacy training for physicians essential before deployment. However, whether structured AI-literacy training translates into improved diagnostic reasoning remains unknown. We conducted a single-blind randomized controlled trial involving 60 licensed physicians from multiple medical institutions in Pakistan, a LMIC, between January 10, 2025, and May 17, 2025. Participants completed a novel 20-hour AI-literacy curriculum covering LLM capabilities, limitations, and appropriate use. Post-training, physicians were randomized to either LLM access plus conventional resources or conventional resources only, with 75 minutes allocated to review up to 6 clinical vignettes. The primary outcome was diagnostic reasoning score (percentage) from a validated, expert-graded rubric assessing differential diagnosis, supporting/opposing factor appropriateness, and next steps; secondary outcome was time per vignette (seconds). Of 58 physicians completing the study, those with LLM access achieved mean diagnostic reasoning scores of 71.4% versus 42.6% with conventional medical resources alone, yielding an adjusted difference of 27.5 percentage points (95% CI, 22.8 to 32.2; P < 0.001). Mean time per case was similar between groups (603.8 vs. 635 seconds; adjusted difference -6.4 seconds, 95% CI -68.2 to 55.3; P = 0.84). While LLM alone outperformed the trained physician group by 11.5 percentage points (95% CI, 5.5 to 17.5; P < 0.001), in 38% of cases the physician plus LLM group surpassed the median LLM-alone performance, highlighting physician-AI complementarities. This trial suggests that AI-literacy training can enable physicians in resourcelimited settings to effectively leverage LLMs for enhanced diagnostic reasoning to address diagnostic gaps in LMICs (ClinicalTrials.gov: NCT06774612).

## Introduction

Diagnostic errors remain a pervasive yet preventable source of patient harm, occurring more frequently in low- and middle-income countries (LMICs).^1-4^ These errors stem from a combination of both cognitive factors (e.g., premature closure and availability bias) and systemic issues (e.g., limited information, time pressure, resource constraints).^5,6^ Addressing them requires multifaceted approaches to improve clinical reasoning and enhance patient safety. Recent advances in Large Language Models (LLMs) have shown promise in diagnostic reasoning and offer potential to bridge diagnostic gaps in resource-constrained settings.^7-13^ However, LLMs can also generate confident yet inaccurate outputs—called “hallucinations” —creating additional safety risks.^14^ Compounding this risk, most current AI implementations in healthcare still lack structured physician AI-literacy training, which represents a critical gap in literature and practice. Safe integration of LLMs into clinical practice requires not only careful implementation and oversight, but also comprehensive AI-literacy training that enables physicians to effectively leverage LLMs while understanding their capabilities and limitations.^15^

Recent studies offer mixed evidence on the effectiveness of LLMs as diagnostic aids, with inconsistent outcomes likely stemming from variations in how physicians are prepared to use them. For instance, a recent trial reported that providing GPT-4 as a diagnostic support tool to physicians, with no prior AI-literacy training, did not improve diagnostic reasoning compared to conventional resources.^9^ Conversely, another study showed that GPT-4 assistance enhanced physician performance on open-ended management reasoning tasks.^8^ Crucially, existing evaluations typically assess AI tools with minimal user preparation, overlooking the learning curve needed for effective human-AI collaboration. This knowledge gap is particularly pronounced in LMICs, where AI could have a transformative impact on healthcare delivery but face unique implementation barriers. These include inadequate technical infrastructure, poor local adaptation of AI tools to existing workflows, and data scarcity.^14,16^ This underscores the need for rigorous, context-specific evaluations that integrate LLM deployment with structured AI-literacy training for clinicians, especially in LMIC settings. Without such evidence, large-scale deployment risks not only tool under-utilization and patient harm but also perpetuating global health inequities through poorly adapted technological solutions that fail to address the unique challenges faced by healthcare systems in resource-limited settings.

This RCT aims to address this knowledge gap by evaluating whether physicians in a resource-limited setting, after receiving structured AI-literacy training, demonstrate better diagnostic performance when given access to a commercial LLM chatbot (ChatGPT 4o [GPT-4o]; OpenAI) plus conventional resources compared to peers who rely solely on conventional diagnostic resources such as online medical databases and Google Search without AI features. Before the RCT, all participants completed a 20-hour AI-literacy training program covering LLM capabilities, prompt engineering techniques, and critical output evaluation strategies. During the trial, participants provided differential diagnoses with supporting and opposing evidence and proposed next steps for each clinical case. Each response was then scored by blinded expert reviewers using the validated rubric. Building on prior research, we adopted a validated structured diagnostic reasoning assessment methodology that provides more meaningful insights into clinical decision-making than traditional assessment methods.^9^

## Results

Sixty licensed physicians were recruited between January 10, 2025, and May 17, 2025. Of these, 58 completed the study (2 withdrew consent during the study period). 29 participants completed the intervention and 29 completed the control as shown in Figure 1. Overall, 35 participants (60%) engaged in virtual encounters, while 23 (40%) participated in person. Median (IQR) years in practice was 8.5 (3.3 to 12 years). Further information on participants is included in Table 1.

**Table 1.** Baseline Participant Characteristics.

| Participant Characteristic | Participants, No. (%) |  |  | P value |
| --- | --- | --- | --- | --- |
|  | Overall (n=58) | Physicians plus conventional resources (n=29) | Physicians plus LLM (n=29) |  |
| **Gender** |  |  |  |  |
| Male | 19 (33) | 8 (28) | 11 (38) | 0.58 |
| Female | 39 (67) | 21 (72) | 18 (62) |  |
| **Specialty** |  |  |  |  |
| Primary Care<sup>a</sup> | 19 (32.8) | 9 (31.0) | 10 (34.5) | 0.7 |
| Medical Subspecialty<sup>b</sup> | 6 (10.3) | 4 (13.8) | 2 (6.9) |  |
| Surgical Subspecialty<sup>c</sup> | 20 (34.5) | 8 (27.6) | 12 (41.4) |  |
| Diagnostic/Support Specialties<sup>d</sup> | 7 (12.1) | 4 (13.8) | 3 (10.3) |  |
| Other/Non-Clinical<sup>e</sup> | 6 (10.3) | 4 (13.8) | 2 (6.9) |  |
| **Years in practice** |  |  |  |  |
| Mean (SD) | 8.4 (5.7) | 8.6 (6) | 8.1 (5.5) | 0.77 |
| Median (IQR) | 8.5 (3.3-12) | 9.0 (3-13) | 8.0 (4-11) | 0.69 |
| **LLM experience** |  |  |  |  |
| I've never used it before (outside of training) | 2 (3) | 1 (3) | 1 (3) | 0.41 |
| I use it rarely (less than once per month) | 7 (12) | 6 (21) | 1 (3) |  |
| I use it occasionally (more than once per month but less than weekly) | 19 (33) | 10 (34) | 9 (31) |  |
| I use it frequently (weekly or more) | 30 (52) | 12 (41) | 18 (62) |  |
| Notes: Percentages in parentheses may not sum to 100% due to rounding.<br><sup>a</sup> Includes family medicine/general practice, internal medicine, pediatrics, and community health officers.<br><sup>b</sup> Includes emergency medicine, gastroenterology, cardiology, nephrology, pulmonology, and acute medicine/infectious diseases.<br><sup>c</sup> Includes general surgery, gynecology/obstetrics, neurosurgery, urology, ophthalmology, oral & maxillofacial surgery, hepato-pancreato-biliary surgery, and breast surgery.<br><sup>d</sup> Includes radiology, pathology, and anesthesiology.<br><sup>e</sup> Includes community medicine/public health, research positions, and house officers/training posts. |  |  |  |  |

**Figure 1:**
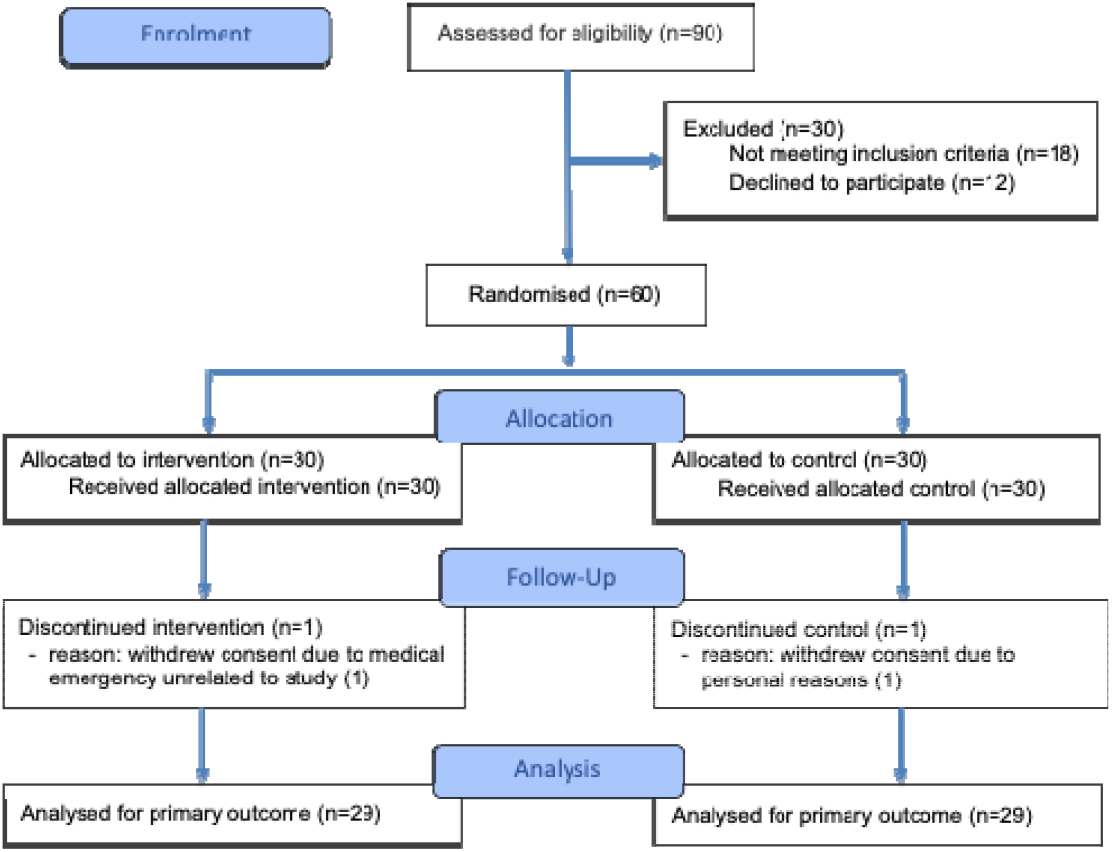
Study flow diagram. The study included 60 practicing attending physicians. Six expert-developed cases were presented, with scoring rubrics created by a panel of three licensed physicians with expertise in clinical reasoning assessment. Physicians were randomized to use either GPT-4o in addition to conventional diagnostic resources (e.g., PubMed, Google Search without AI features), or conventional resources alone. The pre-specified primary outcome was the difference in diagnostic reasoning score between groups on expert-developed scoring rubrics. The pre-specified secondary outcome was the time spent per case.

### Primary Outcome

A total of 342 cases were completed by all participants (172 in the LLM group, and 170 in the control group). The median number of completed cases was 6 (IQR 5-6). The mean (SD) score per case was 71.5% (21.6%) for the LLM group and 42.5% (22.7%) for the control group. The mixed effects model showed a difference of 27.7 percentage points (95% CI: 22.9 to 32.5 pp; *P* < .001) between the LLM and control groups (Table 2 and Figure 2). A sensitivity analysis including all cases, complete and incomplete, showed a similar result with a difference of 28 percentage points (95% CI, 23.2 to 32.8 pp; *P* < .001) between the treatment and control group.

**Table 2.**
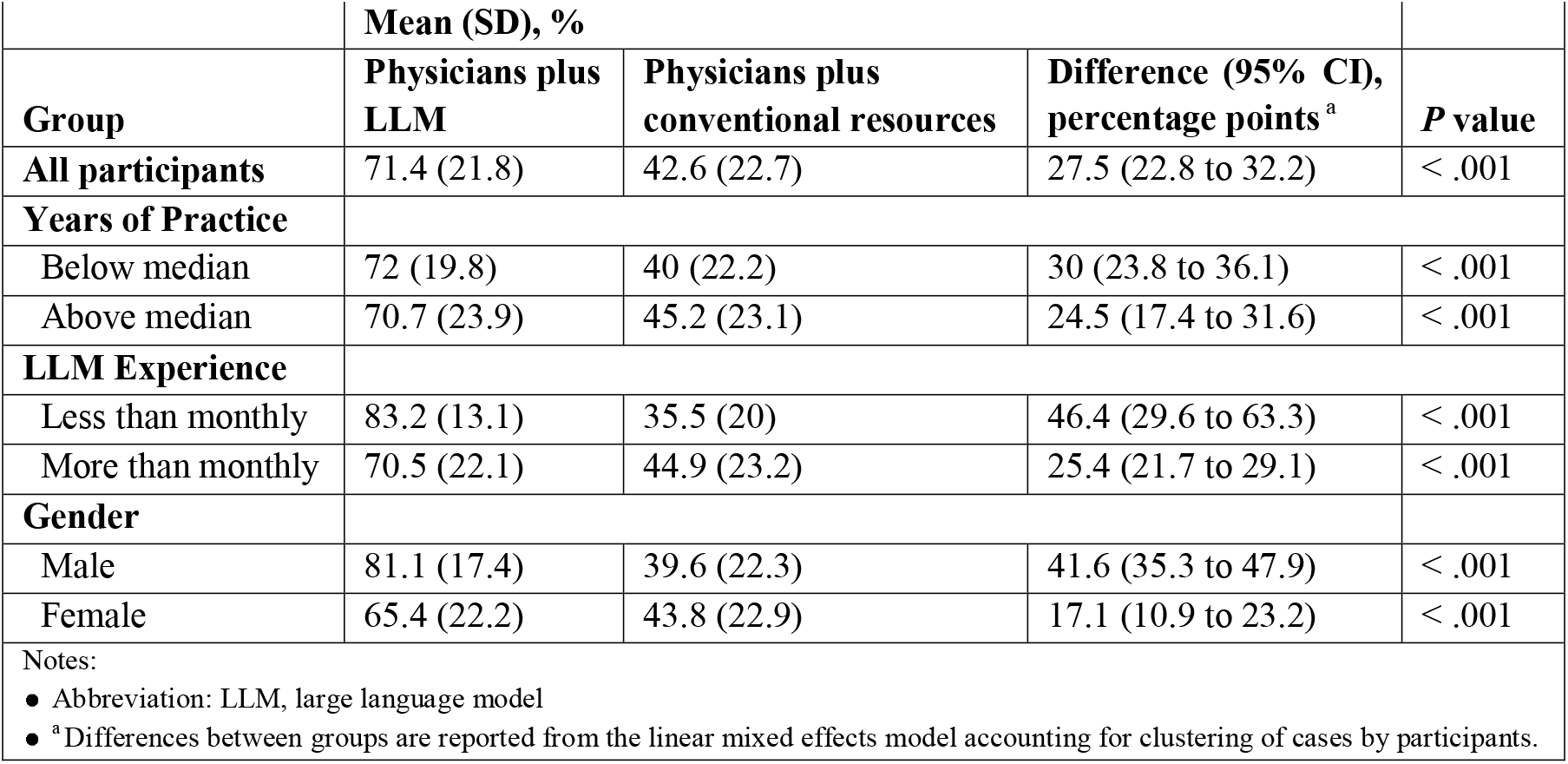
Diagnostic Performance Outcomes.

|  | Mean (SD), % |  |  |  |
| --- | --- | --- | --- | --- |
| Group | Physicians plus LLM | Physicians plus conventional resources | Difference (95% CI), percentage points<sup>a</sup> | P value |
| All participants | 71.4 (21.8) | 42.6 (22.7) | 27.5 (22.8 to 32.2) | < .001 |
| **Years of Practice** |  |  |  |  |
| Below median | 72 (19.8) | 40 (22.2) | 30 (23.8 to 36.1) | < .001 |
| Above median | 70.7 (23.9) | 45.2 (23.1) | 24.5 (17.4 to 31.6) | < .001 |
| **LLM Experience** |  |  |  |  |
| Less than monthly | 83.2 (13.1) | 35.5 (20) | 46.4 (29.6 to 63.3) | < .001 |
| More than monthly | 70.5 (22.1) | 44.9 (23.2) | 25.4 (21.7 to 29.1) | < .001 |
| **Gender** |  |  |  |  |
| Male | 81.1 (17.4) | 39.6 (22.3) | 41.6 (35.3 to 47.9) | < .001 |
| Female | 65.4 (22.2) | 43.8 (22.9) | 17.1 (10.9 to 23.2) | < .001 |
| Notes: |  |  |  |  |
| ● Abbreviation: LLM, large language model<br>● <sup>a</sup> Differences between groups are reported from the linear mixed effects model accounting for clustering of cases by participants. |  |  |  |  |

**Figure 2:**
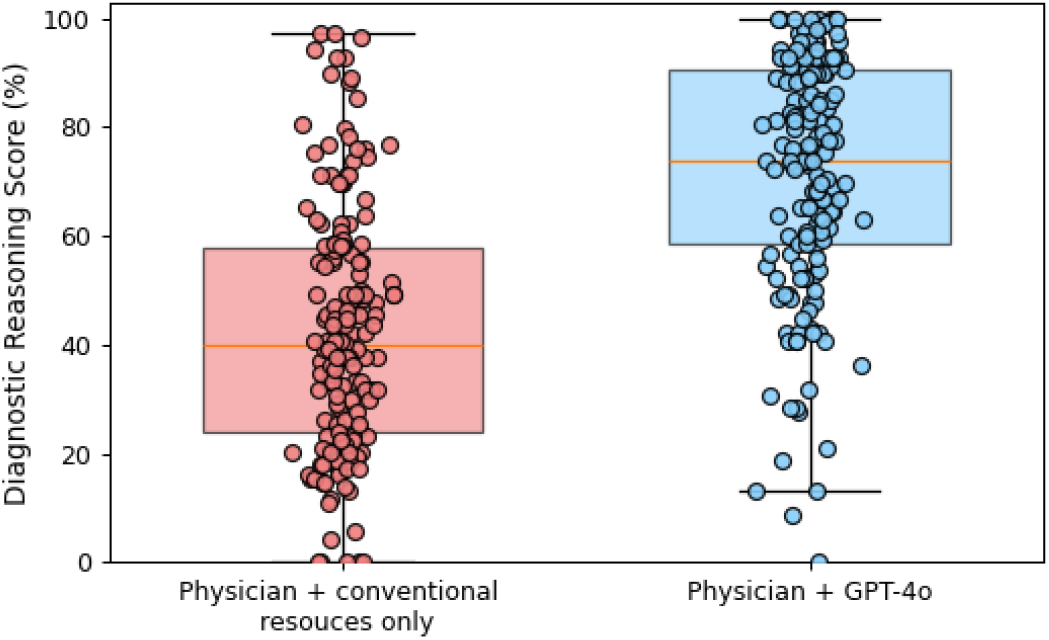
Comparison of the primary outcome for physicians with LLM and with conventional resources only (diagnostic reasoning score standardized to 0–100). Fifty eight physicians (29 randomized to the LLM group and 29 randomized to conventional resources) completed 342 cases (172 in the LLM group, 170 in the conventional resources group). The center of the box plot represents the median, with the boundaries representing the first and third quartiles. The whiskers represent the furthest data points from the center within 1.5 times the IQR.

### Secondary Outcomes

The mean (SD) time spent per case was 603.8 (294.6) seconds for the LLM group and 635 (289.2) seconds for the control group (Figure 3, Table 3). The linear mixed-effects model resulted in an adjusted difference of −6.4 seconds (95% CI, −68.2 to 55.3 seconds; *P* = .84). Analysis of the final diagnosis showed a difference of 34.4 percentage points (95% CI, 31 to 37.9 pp; *P* < .001) between the treatment and control groups (Supplementary Table 1).

**Table 3.** Time Spent per Case.

|  | Mean (SD) time, s |  |  |  |
| --- | --- | --- | --- | --- |
| Group | Physicians plus LLM | Physicians plus conventional resources | Difference (95% CI)<sup>a</sup> | P value |
| All participants | 603.8 (294.6) | 635.0 (289.2) | -6.4 (-68.2 to 55.3) | 0.84 |
| **Years of Practice** |  |  |  |  |
| Below median | 593.2 (295.1) | 640.8 (290.1) | -26.3 (-108.1 to 55.6) | 0.53 |
| Above median | 615.4 (295.4) | 629.1 (289.9) | -11.3 (-100.3 to 77.6) | 0.80 |
| **LLM Experience** |  |  |  |  |
| Less than monthly | 616.4 (210.2) | 709.5 (262.0) | -112.5 (-298.4 to 73.4) | 0.24 |
| More than monthly | 602.8 (300.4) | 610.6 (294.5) | -3.9 (-55 to 47.2) | 0.88 |
| **Gender** |  |  |  |  |
| Male | 582.1 (260.5) | 529.5 (312.4) | 29.6 (-64.1 to 123.4) | 0.54 |
| Female | 617.3 (314.4) | 676.5 (269.8) | -56 (-138.2 to 26.2) | 0.18 |
| Notes: |  |  |  |  |
| ● Abbreviation: LLM, large language model<br>● <sup>a</sup> Differences between groups are reported from the linear mixed effects model accounting for clustering of cases by participants. |  |  |  |  |

**Figure 3:**
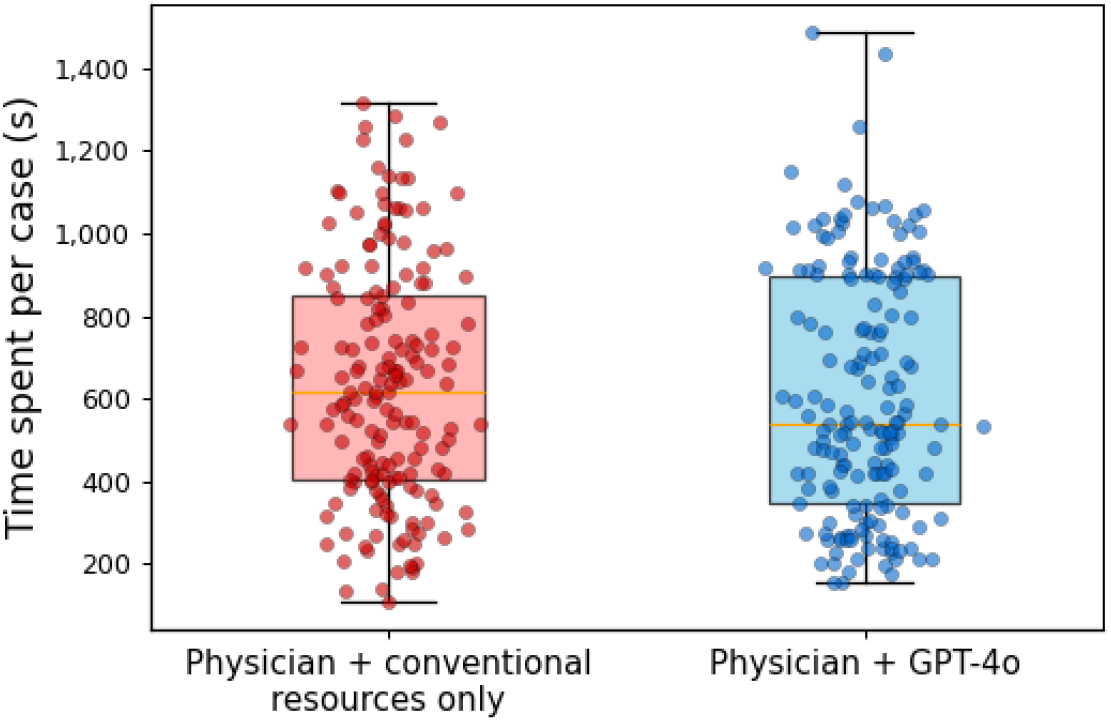
Comparison of the time spent per case by physicians using GPT-4o and physicians using conventional resources only. Fifty-eight physicians (29 randomized to the LLM group and 29 randomized to the conventional resources) completed 342 cases (172 in the LLM group, 170 in the conventional resources group). The center of the boxplot represents the median, with the boundaries representing the first and third quartiles. The whiskers represent the furthest data points from the center within 1.5 times the IQR.

### Subgroup Analyses

Tables 2 and 3 present treatment-versus-control comparisons stratified by three physician characteristics: (1) years of clinical practice since MBBS, (2) self-reported LLM experience, and (3) gender. We find that the treatment effect was larger among those at or below the median years of practice (8.5 years), who experienced a 30 percentage points improvement (95% CI, 23.8 to 36.1 pp; *P* < .001), compared to a 24.5 percentage points improvement among more experienced physicians (95% CI, 17.4 to 31.6 pp; *P* < .001). Similarly, the treatment effect was substantially greater among physicians with less-than-monthly LLM usage experience, who showed a 46.4 percentage points increase in diagnostic accuracy (95% CI, 29.6 to 63.3 pp; *P* < .001), versus a 25.4 percentage point improvement among those using LLMs monthly or less often (95% CI, 21.7 to 29.1 pp; *P* < .001). The treatment benefit also varied significantly by gender, with male physicians experiencing a 41.6 percentage points improvement (95% CI, 35.3 to 47.9 pp; *P* < .001) compared to a 17.1 percentage points improvement among female physicians (95% CI 10.9 to 23.2 pp; *P* < .001).

### LLM Alone

In the three runs of the LLM alone, the mean (SD) score per case was 82.9% (10.4%). Comparing LLM alone with the treatment group found an absolute score difference of 11.5% percentage points (95% CI, 5.5 to 17.5 pp; P = < .001) favoring the LLM alone (Figure 4). Yet, in 38% of cases, the physician plus LLM group surpassed the median performance of the LLM alone group.

**Figure 4:**
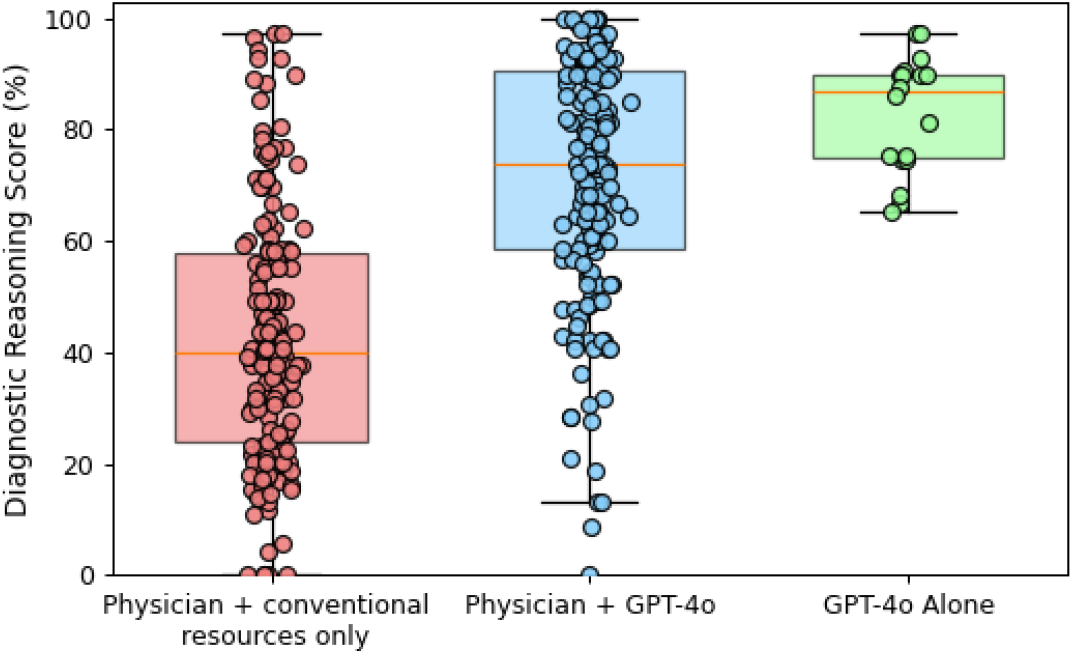
Comparison of the primary outcome according to GPT-4o alone versus physician with GPT-4o and with conventional resources only (total score standardized to 0–100). The GPT-4o alone arm represents the model being prompted by the study team to complete thesix cases, with the models prompted three times for each case for a total of 18 observations. The physicians with GPT-4o group included 29 participants that completed 172 cases, while the physician with conventional resources group included 29 participants that completed 170 cases. The center of the box plot represents the median, with the boundaries representing the first and third quartiles. The whiskers represent the furthest data points from the center within 1.5 times the IQR.

### Assessment Tool Validation

Inter-rater reliability among three graders was high (Krippendorff’s α = 0.85), consistent with diagnostic performance studies, and internal consistency of the grading instrument was strong (Cronbach’s α = 0.8). Inter-rate reliability metrics and variances for individual rubric sections are presented in Supplementary Tables 2 and 3.

## Discussion

This RCT conducted in a LMIC found that AI-trained physicians’ use of GPT-4o, a commercially available chatbot, significantly improved diagnostic reasoning on challenging clinical cases with the largest gains seen in less experienced clinicians, frequent users, and male physicians. All participating physicians completed structured AI-literacy training on LLMs’ capabilities, limitations (including hallucinations), appropriate use, and the necessity of output verification. The training protocol excluded practice with similar clinical vignettes to prevent “teaching to the test” effects and better approximate real-world implementation. Although physicians received instruction in effective LLM prompting techniques, the study design encouraged them to interact naturally with the AI to reflect typical clinical workflows.

Our study presents a notable contrast to earlier research where physicians, without AI-literacy training, did not experience improved diagnostic reasoning from GPT-4 access compared to using conventional resources alone.^9^ In another recent RCT, GPT-4 assistance improved physician performance on open-ended management reasoning tasks, which are related yet significantly different from diagnostic reasoning and involve decision-making around treatment, testing, patient preferences, and risk management.^8^ Considering prior research alongside our findings, underscores the importance of adequate AI-literacy training when integrating advanced AI tools like GPT-4o into clinical practice. Training ensures that healthcare providers can effectively harness the capabilities of LLMs, leading to improved patient outcomes and enhanced clinical decision-making.

Our control group’s diagnostic reasoning score (42.6%) was far below the 74% observed in a U.S. RCT,^9^ suggesting diagnostic reasoning gaps in resource-limited settings. This aligns with WHO data indicating higher diagnostic errors in LMICs.^3^ Such disparities suggest that AI-assisted diagnostic tools could have a large impact in bridging diagnostic gaps in resource-constrained settings. Our findings emphasize that optimizing this synergy requires AI-literacy training of clinicians. The AI-only arm scored 82.9%, highlighting both its promise and the need for human-AI collaboration. In 38% of cases, the physician plus LLM group surpassed the median performance of the LLM alone group, highlighting potential physician-AI complementarities. While LLMs faltered in cases requiring nuanced clinical judgment or subtle red-flag contextual synthesis, physicians with appropriate training could effectively complement these shortcomings. Future research should focus frameworks that leverage the strengths of both human expertise and AI technologies to improve outcomes across diverse healthcare settings.

### Limitations

Our study had several limitations. First, we used clinical vignettes, which were based on, but not actual patient cases. Thus, they may not fully capture real-world clinical decision complexities. Second, we assessed only GPT-4o, chosen for its widespread commercial use medical integration. This limits the generalizability of our findings as it does not capture how alternative LLMs might perform under similar conditions. Third, although we aimed to replicate clinical usage by permitting flexible GPT-4o use, we did not evaluate the need for periodic reinforcement training. Over time, physicians may need refresher sessions to maintain safe, effective AI interaction, especially as models or guidelines evolve.

Finally, our evaluation included six vignettes, limited by session time but comparable to standard medical exams. However, this limited sample cannot capture all clinical scenarios. The selected cases provided broad coverage, but future research should involve a larger array of scenarios and longer observation periods to verify performance consistency and examine clinician adaptation to AI recommendations across varied contexts.

Our randomized clinical trial showed that LLM availability significantly boosts diagnostic reasoning in AI-trained physicians versus those with conventional resources alone, underscoring the need for clinician AI training. While LLMs alone outperformed physicians, a considerable gap persists, highlighting the necessity for frameworks that synergize human expertise with AI in clinical decision support.

## Methods

This study was reviewed and approved by the institutional review board at Lahore University of Management Sciences (LUMS). Informed consent was obtained from all participants prior to enrollment and randomization. Participants received a 1-month free subscription to ChatGPT Plus (worth USD 20) as compensation for completing the study. The study design and reporting adhere to the Consolidated Standards of Reporting Trials (CONSORT) 2025 guidelines for randomized clinical trials. The complete study protocol is available in Supplementary Information 1.

Eligible participants included registered medical practitioners with the Pakistan Medical and Dental Council who possessed a Bachelor of Medicine, Bachelor of Surgery (MBBS) degree and had completed 20 hours of structured AI and LLM training (Supplementary Table 4). Participants were recruited through email distribution lists at the LUMS Learning Institute, which offers specialized training programs for physicians on health AI and data science. Our study spans six months, as participants were recruited from two consecutive cohorts of the training program. Participants were organized into small groups and supervised by study coordinators in either remote sessions or at an in-person computer laboratory at LUMS. Each session lasted 85 minutes, comprising a 10-minute baseline survey followed by 75 minutes dedicated to completing clinical vignette assessments. The trial was completed as planned. Figure 1 illustrates the participant flow, with a visual representation provided in Supplementary Figure 2.

### Clinical Vignettes

Clinical vignettes were adapted from previous studies that set the standard for the evaluation of computer-based diagnostic systems.^11,17^ This study utilized six clinical vignettes based on actual patient cases, which were carefully modified for research purposes. These cases represented moderate diagnostic complexity, excluding both trivial and highly unusual conditions to ensure relevant clinical challenge.^18^ To preserve the integrity of the assessment materials for future administrations, the complete set of clinical vignettes have never been publicly released, and are therefore excluded from the LLM training data corpus; a representative case is included in Supplementary Table 5. Each vignette maintained a consistent format presenting the chief complaint, history of present illness, relevant past medical history, physical examination findings, and laboratory results. This systematic format ensured uniform presentation of clinical information across all cases.

A panel of three physicians among the study co-authors (M.A.K, M.J.A., and A.Z.S.) developed selection criteria for the vignettes. These criteria emphasized appropriate diagnostic complexity, representation across common medical specialties, and clinical relevance, while systematically excluding both diagnostically straightforward and exceptionally rare conditions. The panel reviewed 15 cases, selecting six that met these specifications. Pilot testing demonstrated that most participants completed these cases within 75 minutes, which established the study’s time limit for evaluating vignettes. The cases underwent revision to reflect current laboratory reporting standards and replace specialized medical terminology with descriptive language (e.g., changing “erythema nodosum” to “painful, red, raised bumps on shins”).

While differential diagnosis accuracy is a standard metric in clinical decision support research, such measures inadequately capture the complexity of diagnostic reasoning.^7^ Though we included final differential accuracy as a secondary exploratory measure for consistency with literature, our primary assessment framework used a more nuanced approach to evaluate diagnostic reasoning quality.^9^ We adopted a structured reflection methodology requiring participants to document diagnostic hypotheses with supporting and opposing evidence, reflecting clinical documentation practices. Participants used a template (Supplementary Table 5) to provide their top three differential diagnoses with supporting and contradicting factors, final diagnosis, and up to three recommended next steps.

### Assessing Diagnostic Performance

Our scoring methodology involved the evaluation of the complete assessment grid submitted by participants. Each clinically plausible diagnosis earned up to 1 point based on its relevance and likelihood. Supporting and opposing findings identified by participants received 0-2 points according to correctness (0 for incorrect or missing evidence, 1 for partially correct or incomplete evidence, and 2 for fully correct and comprehensive evidence). Final diagnoses were awarded 2 points for the most accurate diagnosis and 1 point for clinically plausible or insufficiently specific responses, while incorrect diagnoses received no points. Finally, participants provided up to 3 next steps to further evaluate the patient with 1 point awarded for a partially correct response and 2 points for a completely correct response based on their clinical appropriateness (detailed scoring criteria available in Supplementary Tables 6 and 7). Incorrect diagnoses earned no points. This scoring approach provided assessment of both diagnostic accuracy and clinical reasoning quality. To ensure objectivity, evaluators were blinded to participant group assignments by receiving all responses in an identical structured reflection template, with timestamps and other identifying metadata removed, preventing them from distinguishing between responses generated using LLM assistance versus conventional resources.

### Study Design

We employed a randomized single-blind study design. Participants were assigned to either the intervention group (access to GPT-4o) or the control group (conventional resources only). Both groups could utilize conventional medical resources (e.g., online medical databases, Google Search without AI features), while the intervention group could additionally use GPT-4o. Control group participants were prohibited from using LLMs, and a browser extension blocked Google ‘AI Overviews.’ No concomitant care was applicable as participants were physicians completing assessments, not patients. Study accounts were provided for LLM access, with interactions recorded. Participants had 75 minutes to complete up to 6 diagnostic cases, with emphasis placed on response quality over quantity.

The study was implemented using a secure survey platform (Kobotoolbox), with clinical vignettes presented in randomized order to each participant to minimize potential case-order bias. Because the intervention was judged a priori to pose no more than minimal risk, we did not prespecify individual adverse-event categories. Harms were therefore assessed non-systematically by recording withdrawals and incidents during test sessions.

In addition to our primary analysis, we conducted a secondary comparison with an LLM-only arm. Following established prompt engineering principles, we developed a zero-shot prompt through iterative refinement; this prompt was consistently applied across all clinical vignettes without modification. A researcher administered these prompts to the model and documented responses (example prompt provided in Supplementary Table 8). To ensure robust evaluation of the model’s performance, we conducted three independent runs in separate sessions, with all outputs included in the blinded grading process alongside human participant responses. This approach ensured that all LLM-generated assessments underwent identical evaluation procedures as human outputs.

### Validation of Assessment Tool

To finalize the assessment rubrics, three licensed physicians with expertise in clinical reasoning assessment (M.A.K, M.J.A., and A.Z.S) independently solved each clinical vignette to establish inter-rater reliability and flag potential scoring discrepancies. Disagreements were resolved through structured consensus discussions, resulting in standardized scoring rubrics for each case that captured essential diagnostic elements while acknowledging legitimate clinical variations. Each participant’s responses underwent blinded evaluation by three qualified scorers; componentwise scoring variance appears in Supplementary Table 3. The arithmetic mean of the three ratings constituted the diagnostic reasoning score for each case. Our methodology deliberately accommodated the inherent ambiguity in clinical diagnosis by allowing multiple correct response variations when supported by scorer consensus. Statistical validation included Krippendorff’s alpha for inter-rater reliability and Cronbach’s alpha for the instrument’s internal consistency.

### Study Outcomes

Our prespecified primary outcome was the composite diagnostic reasoning score, calculated as a percentage of total possible points across all components of the structured reflection assessment tool. Our prespecified secondary outcome was time spent per case (measured in seconds). Final diagnosis accuracy was assessed as an exploratory outcome. All outcomes were compared at the case-level between the randomized groups.

### Statistical Analysis

We summarized outcomes with descriptive statistics and compared demographic characteristics between groups using χ^2^ test or Fisher exact test for categorical variables and two-sided *t* test or Mann-Whitney *U* test for the mean and median of continuous variables, respectively.

A target sample size of 50 participants (25 per arm) was predetermined based on a prior study.^9^ Ultimately, 60 participants enrolled, and 58 completed the study. Power analysis, conducted using Python version 3.11.9 (Python Software Foundation) and statsmodels version 0.14.4 (Statsmodels Developers), indicated that a sample of 200 completed cases (4 per participant) would provide at least 80% power to detect an 8-percentage-point mean difference in scores, assuming a two-sided α of .05. The analysis employed mixed-effects models suitable for cluster-randomized designs, considering an intraclass correlation coefficient ranging from 0.05 to 0.15 and standard deviation of 16.2%.

All analyses followed the intention-to-treat principle and were performed at the case level, with cases clustered by participants. Linear mixed-effects models evaluated differences in primary and secondary outcomes, both treated as continuous variables, with verified normality assumptions (Supplementary Figure 1). Random effects were included for participants to account for within-participant correlations and for cases to control for case difficulty variability.

Family-wise error rate was maintained at α = 0.05 using the Holm-Bonferroni method for the prespecified primary outcome. Secondary outcome analyses were exploratory and did not include adjustments for multiple comparisons. Final diagnosis accuracy was likewise evaluated post hoc without adjustment for multiple comparisons. Preplanned sensitivity analysis assessed the impact of incorporating incomplete cases on the primary outcome. Subgroup analyses were performed based on years of practice post-MBBS, prior LLM experience, and gender. An additional exploratory analysis treated LLM-only attempts as a separate group in a nested design (three attempts per participant) and compared them with human-completed cases.

All statistical analyses used Python version 3.11.12 (Python Software Foundation) with pandas for data manipulation and statsmodels version 0.14.4 for mixed-effects modeling.

## Data Availability

Example case vignettes, questions, and the scoring rubric are provided in the Supplementary File. Deidentified participant-level data is available for non-commercial purposes upon reasonable request and contingent on a signed data-use agreement. Data requests should be directed to the corresponding author (I.A.Q) at.

## Code Availability

Python scripts enabling the main steps of the analysis are available from the corresponding author upon reasonable request.

## Acknowledgment

This work was supported by an internal Faculty Initiative Fund grant (FIF-0912) and the Learning Institute at Lahore University of Management Sciences (LUMS). We thank Ushna Malik and Laiba Intizar Ahmad for administering the surveys and proctoring study sessions; each served as a paid research assistant on this project and received compensation from study funds.

## Author Contributions

I.A.Q., A.A., A.U.K., M.J.A., A.Z.S., and M.H.A. contributed to concept and design. I.A.Q., A.A., A.U.K., A.Z.S., M.J.A., and M.H.A. contributed to acquisition, analysis, or interpretation of data. I.A.Q. and A.A. contributed to drafting of the manuscript. I.A.Q. and A.A. contributed to statistical analysis. I.A.Q., A.A., and M.H.A. contributed to obtaining funding. I.A.Q., A.A., A.U.K., A.Z.S., M.J.A., and M.H.A. contributed to administrative, technical, or material support. I.A.Q. and A.A. contributed to supervision. All authors reviewed the manuscript.

## Competing interests

The authors declare no competing interests.

**Correspondence** and requests for materials should be addressed to Ihsan Ayyub Qazi

## Patient and Public Involvement

Not applicable. This study will involve only physicians as participants and aims to evaluate clinical decision-making performance using standardized vignettes. No patients will be recruited as research participants.

