## Supplementary File for "AI-literacy training enhances physician-LLM diagnostic collaboration in a resource-limited setting: a randomized controlled trial"

**Supplementary Materials**

- Supplementary Table 1: Impact of treatment when using final diagnosis as the outcome (exploratory)
- Supplementary Table 2. Inter-Rater Reliability by Component
- Supplementary Table 3. Assessment Tool Validation: Component-wise Variance Between Scores Assigned by Scorers
- Supplementary Table 4. AI Training Topics
- Supplementary Table 5. Diagnostic Case-1 Vignette and Questions
- Supplementary Table 6. Structured Reflection Rubric for Diagnostic Case-1 - High-Scoring Example Response
- Supplementary Table 7. Structured Reflection Rubric for Diagnostic Case-1 - Low-Scoring Example Response
- Supplementary Table 8: GPT-4o Prompt and Responses for Diagnostic Case-1
- Supplementary Figure 1. Statistical Validation of Normality
- Supplementary Figure 2. Study Flow Diagram
- Supplementary Information 1: Study Protocol
- Supplementary Information 2: Statistical Analysis Plan

**Supplementary Table 1: Impact of treatment when using final diagnosis as the outcome (exploratory)**

| **Final Diagnosis Performance Outcome** | | | | |
| --- | --- | --- | --- | --- |
|  | **Mean (SD), %** | |  |  |
| **Group** | **Physicians plus LLM** | **Physicians plus conventional resources** | **Difference (95% CI), percentage points** ^a^ | ***P* value** |
| **All participants** | 80.8 (36.6) | 46.4 (47.4) | 34.4 (31 to 37.9) | < .0001 |
| Notes:   - Abbreviation: LLM, large language model - ^a^ Differences between groups are reported from the linear mixed effects model accounting for clustering of cases by participants. | | | | |

**Supplementary Table 2. Inter-Rater Reliability by Component**

| **Component** | **Krippendorff's Alpha** |
| --- | --- |
| Diagnoses | 0.994 |
| Support Diagnosis | 0.987 |
| Opposing Diagnosis | 0.981 |
| Final Diagnosis | 0.975 |
| Next Steps | 0.966 |
| Total Marks | 0.852 |

**Supplementary Table 3. Assessment Tool Validation: Component-wise Variance Between Scores Assigned by Scorers**

| **Component** | **Variance** |
| --- | --- |
| Diagnoses | 0.015 |
| Support Diagnosis | 0.076 |
| Opposing Diagnosis | 0.105 |
| Final Diagnosis | 0.052 |
| Next Steps | 0.140 |
| Total Marks | 0.055 |

**Supplementary Table 4. AI Training Topics.**

| **AI Training Topics** | |
| --- | --- |
| 1. | **Introduction to Artificial Intelligence (AI)**   - What is AI and why it matters - AI as a helpful assistant: from mechanical tasks to intelligent support |
| 2. | **Basics of Machine Learning (ML)**   - What is machine learning and how it works - Overview of ML types - Real-life examples, including healthcare applications |
| 3. | **Basics of Generative AI (GenAI) and Prompt Engineering**   - How GenAI works in simple terms - Introduction to prompt engineering techniques |
| 4. | **Writing and Research using GenAI**   - Writing professional and effective emails with AI assistance - Retrieval-Augmented Generation (RAG) for improved outputs |
| 5. | **AI-powered Presentation Design**   - Creating presentations from descriptions and documents - Prompt engineering techniques specific to presentation generation |
| 6. | **Multimedia Content Creation with GenAI**   - Generating and editing images using AI - Creating audio and video content using simple prompts - Digitizing and summarizing medical notes with AI |
| 7. | **GenAI for Data and Math Tasks**   - Using GenAI to explore and understand data - Prompting for mathematical and data analysis tasks |
| 8. | **GenAI Customization and Limitations**   - Using and building your own custom GPTs - Understanding AI limitations like hallucinations and how to manage them |

**Supplementary Table 5. Diagnostic Case #1 Vignette and Questions**

**Diagnostic Vignette**

**History of Present Illness**

A male in his late 50s presents with swelling and pain in his right leg that started 3 days ago. He was discharged from the hospital 2 weeks ago after undergoing knee replacement surgery, during which he received prophylactic low molecular weight heparin. He reports intermittent fever and recent episodes of palpitations. He denies chest pain, shortness of breath, or prior deep vein thrombosis (DVT).

**Past Medical History**

He has a history of osteoarthritis requiring knee replacement. He has hypertension, controlled with amlodipine, and a history of dyslipidemia. He has no history of malignancy or clotting disorders.

**Physical Examination**

- **VITALS**: BP 125/80 mmHg, pulse 102/min, temperature 99.6°F.
- **GEN**: Appears mildly uncomfortable.
- **CARDIAC**: Regular rhythm, no murmurs or gallops.
- **PULM**: Clear to auscultation bilaterally.
- **ABD**: Soft, non-tender.
- **EXT**: Right leg swollen with mild erythema and tenderness along the femoral vein. Homans' sign is positive. No ulcers or skin changes.
- **SKIN**: Petechiae on both forearms.

**Laboratory Results**

Platelet count has decreased to 58 x 10³/μL from 210 x 10³/μL 10 days ago. D-dimer is elevated at 5.2 μg/mL. The Heparin-PF4 antibody test is positive. Ultrasound reveals a thrombus in the right femoral vein.

**PART 1 – Structured Diagnostic Reasoning Grid**
**List three possible diagnoses for the patient's condition and provide supporting and opposing evidence for each to guide differential diagnosis.**

**A. Diagnosis 1:**

a. Supporting Evidence for Diagnosis 1:
 Provide findings/risk factors supporting Diagnosis 1.

b. Opposing Evidence for Diagnosis 1:
 Provide findings opposing Diagnosis 1, or findings that were expected but are not present.

**B. Diagnosis 2:**

a. Supporting Evidence for Diagnosis 2:
 Provide findings/risk factors supporting Diagnosis 2.

b. Opposing Evidence for Diagnosis 2:
 Provide findings opposing Diagnosis 2, or findings that were expected but are not present.

**C. Diagnosis 3:**

a. Supporting Evidence for Diagnosis 3
 Provide findings/risk factors supporting Diagnosis 3.

b. Opposing Evidence for Diagnosis 3:
 Provide findings opposing Diagnosis 3, or findings that were expected but are not present.

**PART 2 - Final Diagnostic Decision:**

Based upon your reasoning above, what is your final diagnosis?

**PART 3 - Additional Steps:**
**List up to 3 additional steps that you would take in your diagnostic process:**

Step 1

Step 2

Step 3

**Supplementary Table 6. Structured Reflection Rubric for Diagnostic Case-1 - High-Scoring Example Response.** Example of a high-scoring response. The participant provided three plausible diagnoses with accurate and comprehensive supporting and opposing evidence and reached the correct final diagnosis.

| **Question** | **High Scoring Example** | **Score** |
| --- | --- | --- |
| **Question 1:**  Diagnosis - List 3 Possible Diagnoses  (1 point each) | 1. Heparin-Induced Thrombocytopenia (HIT)  2. Deep Vein Thrombosis (DVT)  3. Fat embolism | 3/3 |
| **Question 2:**  Support Diagnosis - For each possible diagnosis, provide findings/risk factors supporting this hypothesis  (2 points each) | 1. Heparin-Induced Thrombocytopenia (HIT):  - Recent heparin use (low molecular weight  heparin for knee surgery).  - New-onset thrombosis (right femoral vein  thrombus) and thrombocytopenia (platelet  count decreased).  - Positive Heparin-PF4 antibody test.  - Petechiae and fever are signs of systemic  Involvement.  2. Deep Vein Thrombosis (DVT):  - Swelling and tenderness along the femoral  veins are suggestive of DVT. However, the  associated thrombocytopenia and positive Heparin-PF4 antibody test suggest HIT rather than typical DVT.  3. Fat Embolism:  -Osseous manipulation & petechial rash on  forearms. Short time duration between the onset of petechial rash and knee replacement surgery. | 6/6 |
| **Question 3**  Opposing Diagnosis - For each possible diagnosis, provide findings opposing this hypothesis, or findings that were expected but not present (2 points each) | 1. Heparin-Induced Thrombocytopenia (HIT):  - No opposing findings. The history of recent heparin exposure, thrombocytopenia, positive  Heparin-PF4 antibodies, and new thrombosis  support HIT.  2. Deep Vein Thrombosis (DVT):  - HIT explains the thrombosis better, as DVT typically wouldn't cause thrombocytopenia or be associated with a positive Heparin-PF4 antibody test.  3. Fat Embolism:  - No neurological or respiratory symptoms | 6/6 |
| **Question 4:**  Final Diagnosis  (2 points) | Heparin-Induced Thrombocytopenia (HIT): The clinical presentation of new thrombosis, thrombocytopenia, recent heparin use, positive Heparin-PF4 antibody test, and petechiae confirm HIT. | 2/2 |
| **Question 5:**  Additional Steps  (2 points each) | 1. Discontinue all heparin products immediately to prevent further thrombotic events.  2. Start alternative anticoagulation (e.g., argatroban or fondaparinux) for anticoagulation management.  3. Monitor platelet count closely to assess recovery and prevent further thrombosis. | 6/6 |
| **Total Score** |  | 23/23 |

**Supplementary Table 7. Structured Reflection Rubric for Diagnostic Case-1 - Low-Scoring Example Response.** Example of a low-scoring response. The participant provided only one plausible diagnosis with appropriate supporting and opposing evidence.

| **Question** | **Low Scoring Example** | **Score** |
| --- | --- | --- |
| **Question 1:**  Diagnosis - List 3 Possible Diagnoses  (1 point each) | 1. Deep Vein Thrombosis (DVT)  2. Drug Induced Bleeding  3. Immune Thrombocytopenic Purpura (ITP) | 1/3 |
| **Question 2:**  Support Diagnosis - For each possible diagnosis, provide findings/risk factors supporting this hypothesis  (2 points each) | 1. Deep Vein Thrombosis (DVT):  - Swelling and pain in rt leg, surgical history and ultrasonography findings with D-dimers and Homan's sign  2. Drug Induced Bleeding:  - History of anticoagulation therapy and heparin studies  3. Immune Thrombocytopenic Purpura (ITP):  - Thrombocytopenia with petechiae, purpura and bleeding tendency | 2/6 |
| **Question 3**  Opposing Diagnosis - For each possible diagnosis, provide findings opposing this hypothesis, or findings that were expected but not present  (2 points each) | 1. Deep Vein Thrombosis (DVT):  - Petechiae on forearm and no chest findings  2. Drug Induced Bleeding:  - Ultrasonography leg suggestive of DVT  3. Immune Thrombocytopenic Purpura (ITP):  - Dopplers findings. | 2/6 |
| **Question 4:**  Final Diagnosis  (2 points) | Deep vein thrombosis with thrombocytopenia and consumptive coagulopathy | 0/2 |
| **Question 5:**  Additional Steps  (2 points each) | 1. D-dimers, fibrin degradation product  2. Peripheral film, bone marrow biopsy  3. Doppler USG | 0/6 |
| **Total Score** |  | 5/23 |

**Supplementary Table 8: GPT-4o Prompt and Responses for Diagnostic Case-1**

| **GPT-4o Prompt** | **GPT-4o Response** |
| --- | --- |
| You are a medical doctor with an MBBS degree, who is solving a complex medical case for a test.  You are going to receive a case vignette. After reading the case, I want you to give three parts of information.  Here is the first part: First, you will list three possible diagnoses. Please number these 1., 2., 3.  Next, for each of the three diagnoses listed, you will provide the findings or risk factors that support this hypothesis.  Next, for each of the three diagnoses listed, you will provide findings opposing this hypothesis,  or findings that were expected but not present.  Part 2 – Final diagnostic decision. Here is the second part: Please answer this question: Based upon your reasoning above, what is your final diagnosis?  Part 3 – Additional Steps. Here is the third part: Name up to 3 additional steps you would take in your diagnostic process.  Diagnostic Case Here is the case: | Part 1 - Diagnostic Hypotheses:  1. Deep Vein Thrombosis (DVT) due to Heparin-Induced Thrombocytopenia (HIT)  - Supporting findings/risk factors:  - Recent surgery (knee replacement), leading to immobilization and increased risk of DVT.  - Positive Homans' sign, swelling, erythema, and tenderness along the femoral vein, all indicative of DVT.  - Thrombus identified in the right femoral vein via ultrasound.  - Decrease in platelet count post-surgery, with a significant drop from 210 x 10³/μL to 58 x 10³/μL.  - Positive Heparin-PF4 antibody test, which supports HIT as a cause of thrombocytopenia and thrombosis.    - Opposing findings:  - None. The combination of thrombocytopenia, positive Heparin-PF4 test, thrombus, and recent heparin use aligns well with HIT complicating to DVT.  2. Post-surgical Infection leading to Septic Phlebitis  - Supporting findings/risk factors:  - Intermittent fever and elevated temperature (99.6°F) could suggest infection.  - Swelling, erythema, and tenderness in the right leg could indicate phlebitis.  - History of recent surgery, which can be a risk factor for infection.  - Opposing findings:  - Absence of leukocytosis or other systemic signs of a significant infection.  - Presence of petechiae and significant thrombocytopenia, along with the positive Heparin-PF4 test, is more suggestive of HIT rather than infection.  - The ultrasound finding of a thrombus and the drop in platelets without clear infection markers make infection less likely as the primary cause.  3. Osteomyelitis or Prosthetic Joint Infection  - Supporting findings/risk factors:  - Recent knee replacement surgery with consequent risk of infection.  - Intermittent fever and increased temperature could suggest an infectious process.  - Opposing findings:  - The site of pain and swelling is along the femoral vein, not the knee joint, which decreases the likelihood of a prosthetic joint infection or osteomyelitis.  - Lack of severe joint-specific symptoms or drainage from the surgical site.  - The thrombocytopenia and positive Heparin-PF4 antibody test align more with HIT than a prosthetic joint infection.  Part 2 - Final diagnostic decision:  Based on the reasoning above, the final diagnosis is Deep Vein Thrombosis (DVT) due to Heparin-Induced Thrombocytopenia (HIT).  Part 3 - Additional Steps:  1. Immediately discontinue all heparin products and avoid further use.  2. Initiate alternative anticoagulation with a non-heparin anticoagulant, such as a direct thrombin inhibitor (e.g., argatroban) or a factor Xa inhibitor.  3. Monitor platelet counts, coagulation parameters, and thrombus resolution closely while managing the HIT and associated DVT. |

**Supplementary Figure 1. Statistical Validation of Normality.** Residual-distribution diagnostics for the mixed effects model. Panels show (top-left) a QQ plot of the model residuals against a theoretical normal distribution; (top-right) a histogram of residuals; (bottom-left) a kernel density estimate (shaded) of the residuals overlaid with the corresponding normal distribution curve (red line; mean = 0, SD = 2.51); and (bottom-right) results of the Shapiro–Wilk test for normality, including W statistic, P value, and interpretation of the null hypothesis that the data are normally distributed.

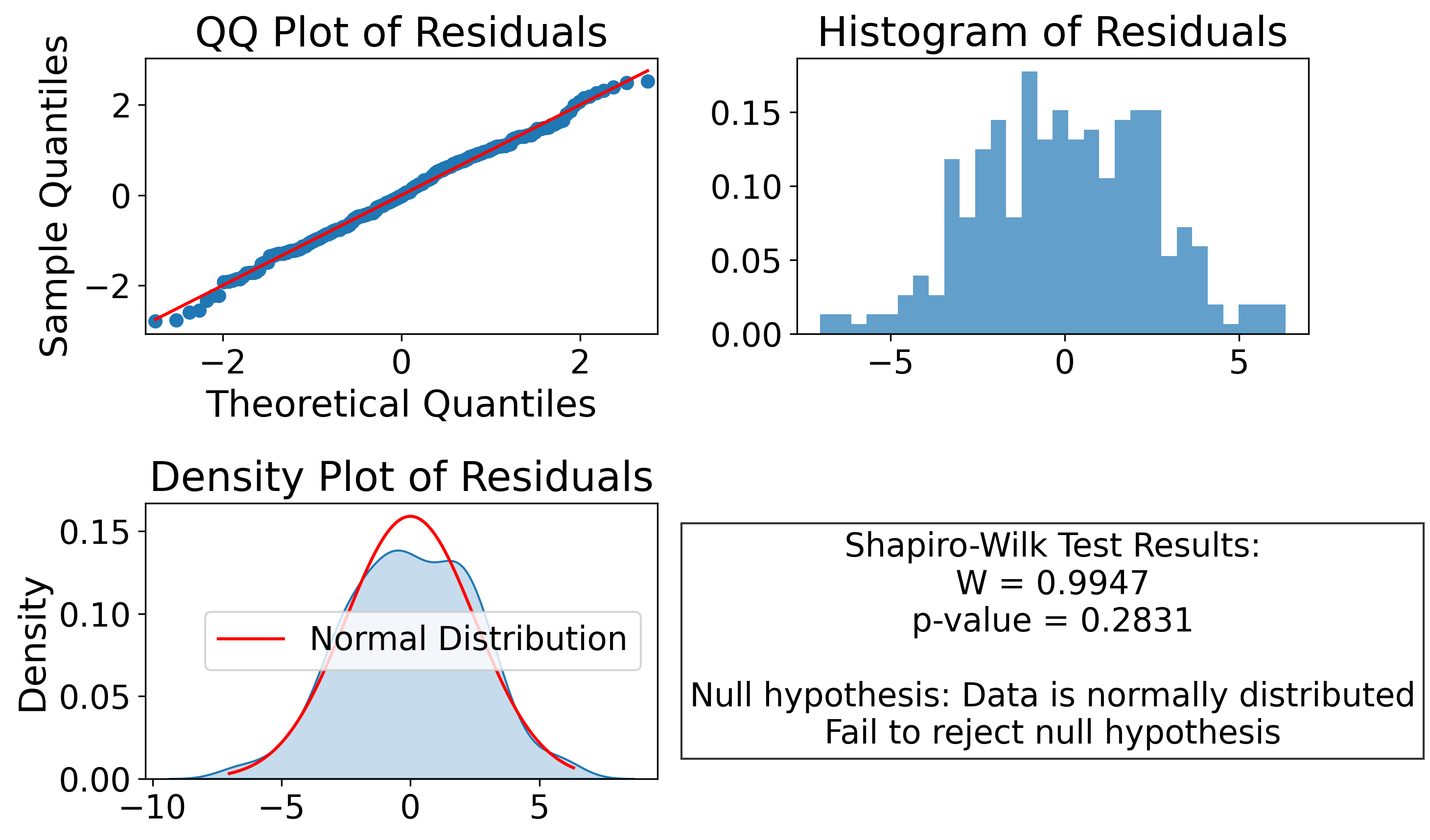

**Supplementary Figure 2. Study Flow Diagram.** Sixty physicians were randomized to complete diagnostic vignettes using either GPT-4o and conventional resources or conventional resources alone. Participants provided differential diagnoses with supporting evidence for and against each diagnosis and recommended optimal next diagnostic evaluation steps.

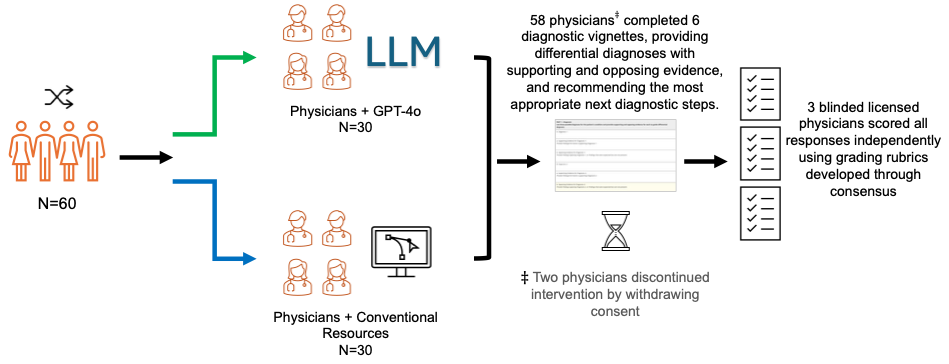

### Supplementary Information 1. Study Protocol

This study was prospectively registered at ClinicalTrials.gov prior to data collection ([NCT04011644](https://clinicaltrials.gov/study/NCT06774612)).

**Brief Summary**: This study aims to evaluate whether large language model-trained medical doctors demonstrate enhanced diagnostic reasoning performance when utilizing ChatGPT-4o alongside conventional resources compared to using conventional resources alone.

**Condition or disease**: Diagnosis

**Intervention/treatment**: Other: ChatGPT-4o

**Phase**: Not Applicable

### Detailed Description:

Diagnostic errors are a major source of preventable patient harm. Recent advances in Large Language Models (LLM), particularly ChatGPT-4o, have shown promise in enhancing medical decision-making. However, little is known about their impact on medical doctors’ diagnostic reasoning. Diagnostic accuracy relies on complex clinical reasoning and careful evaluation of patient data. While AI assistance could potentially reduce errors and improve efficiency, ChatGPT-4o lacks medical validation and could introduce new risks through incorrect information generation (also known as hallucinations). To mitigate these risks, doctors need adequate training in understanding ChatGPT-4o's capabilities, limitations, and proper usage. Given these uncertainties and the importance of proper AI training, systematic evaluation is essential before clinical implementation.

This randomized study will assess whether ChatGPT-4o access improves LLM-trained medical doctors' diagnostic performance compared to conventional resources (e.g., textbooks, online medical databases) alone. All participating doctors will have completed at least a 10-hour training program covering ChatGPT-4o usage, prompt engineering techniques, and output evaluation strategies. Participants will provide differential diagnoses with supporting evidence and recommended next steps for clinical cases, with responses evaluated by blinded reviewers.

***Study Design***

Study Type: Interventional (Clinical Trial)

Actual Enrollment: 58 participants

Allocation: Randomized. Randomization list was created by Ihsan Ayyub Qazi using the Sealed Envelope program.

Intervention Model: Parallel Assignment

Intervention Model Description: The trial will be designed as a randomized, two-arm, single-blind parallel group study.

Masking: Single (Outcomes Assessor)

Masking Description: The grading of responses will be performed by assessors blinded to participant identity and treatment assignment.

Primary Purpose: Diagnostic

Official Title: Diagnostic Reasoning With and Without AI Support: A Randomized Controlled Trial of LLM-Trained Medical Doctors

Actual Study Start Date: January 10, 2025

Actual Primary Completion Date: May 17, 2025

Actual Study Completion Date: May 17, 2025

***Arms and Interventions***

| **Arm** | **Intervention/Treatment** |
| --- | --- |
| Active Comparator: ChatGPT-4o Group will be given access to ChatGPT-4o. | Other: OpenAI's ChatGPT-4o large language model with chat interface. |
| No Intervention: Group will not be given access to ChatGPT-4o but will be encouraged to use any resources they wish besides large language models (PubMed, Google without AI Overviews, etc). |  |

***Outcome Measures***

**Primary Outcome Measures:**

Diagnostic reasoning [Time Frame: Assessed at a single time point for each case, during the scheduled diagnostic reasoning evaluation session, which takes place between 0-4 days after participant enrollment.]

The primary outcome will be the percent correct for each case (range: 0 to 100). For each case, participants will be asked for three top diagnoses, findings from the case that support that diagnosis, and findings from the case that oppose that diagnosis. For each plausible diagnosis, participants will receive 1 point. Findings supporting the diagnosis and findings opposing the diagnosis will also be graded based on correctness, with 1 point for partially correct and 2 points for completely correct responses. Participants will then be asked to name their top diagnosis, earning one point for a reasonable response and two points for the most correct response. Finally participants will be asked to name up to 3 next steps to further evaluate the patient with one point awarded for a partially correct response and two points for a completely correct response. The primary outcome will be compared on the case-level by the randomized groups.

**Secondary Outcome Measures:**

Time Spent on Diagnosis [Time Frame: Assessed at a single time point for each case, during the scheduled diagnostic reasoning evaluation session, which takes place between 0-4 days after participant enrollment.]

We will compare how much time (in seconds) participants spend per case between the two study arms.

***Eligibility Criteria***

Ages Eligible for Study: All

Sexes Eligible for Study: All

Accepts Healthy Volunteers: Yes

**Criteria**

Inclusion Criteria:

- Full or Provisionally Registered Medical Practitioners with the Pakistan Medical and Dental Council (PMDC).
- Completed Bachelor of Medicine, Bachelor of Surgery (MBBS) Exam. The equivalent degree of MBBS in US and Canada is called Doctor of Medicine (MD).
- Participants must have completed a structured training program on the use of ChatGPT (or a comparable large language model), totaling at least 10 hours of instruction. The program must include hands-on practice related to LLM’s aspects, specifically prompt engineering and content evaluation.

Exclusion Criteria:

- Any other Registered Medical Practitioners (Full or Provisional) with PMDC (e.g., Professionals with Bachelor of Dental Surgery or BDS).

**Potential Risks and Harms**

This study poses minimal risk to participants. Some participants may experience mild discomfort or frustration, particularly those in the control group who may find clinical case studies challenging without large language model assistance.

**Safeguards for Addressing Risks or Harms to Subjects**

To minimize potential discomfort, participants will receive comprehensive instructions and ongoing support throughout the study. Participants will be explicitly informed that individual performance is not being evaluated, and that data will be analyzed for aggregate trends only. All participants will be advised of their right to withdraw from the study at any time without penalty or explanation. Research staff will monitor for signs of distress and provide appropriate support as needed.

***Contacts and Locations***

Locations

Pakistan, Lahore

Lahore University of Management Sciences

Lahore, Punjab 54792

Sponsors and Collaborators

Lahore University of Management Sciences

Investigators

Principal Investigator: Ihsan Ayyub Qazi, PhD, Lahore University of Management Sciences

Principal Investigator: Ayesha Ali, PhD, Lahore University of Management Sciences

Principal Investigator: Muhammad Asadullah Khawaja, MBBS, King Edward Medical University

### Supplementary Information 2. Statistical Analysis Plan

**Power Analysis:**

The minimum target sample size of 50 participants (25 participants per arm) was predetermined based on a prior study.^1^ The power analysis, conducted using Python version 3.11.9 (Python Software Foundation), employed the statsmodels.stats.power module from statsmodels version 0.14.4 (Statsmodels Developers) and indicated that a total sample of 200 to 250 completed cases (approximately 4 to 5 cases per participant) would provide at least 80% power to detect an 8-percentage-point mean difference in diagnostic reasoning scores, assuming a two-sided α of .05. The analysis employed mixed-effects models suitable for cluster-randomized designs, considering an intraclass correlation coefficient (ICC) ranging from 0.05 to 0.15 and a standard deviation of 16.2%.

**Statistical Analysis:**

Descriptive analysis will be performed by comparing participant characteristics between the LLM and non-LLM group. Categorical variables will be reported using counts and proportions, while continuous variables will be reported as means and standard deviations or, if their distribution is non-normal, as medians with interquartile ranges.

The primary outcome analysis will be conducted at the case level, with clustering by participant under an intention-to-treat framework. The primary analysis will include cases with completed responses. We will first summarize the mean and standard deviation of scores (standardized on a 0–100 scale) and report the mean and standard deviation for the time spent on each case, both for the overall cohort and separately for the GPT-4o group compared to the conventional resources-only group. To evaluate the impact of using GPT-4o, we will apply generalized mixed-effect models, including a random effect for the participant to account for potential within-participant correlation between cases, and a random effect for cases to account for differences in difficulty across cases. We will also conduct the following sensitivity analysis:

1) Include incomplete cases on the primary outcome.
2) Include past experience in LLM use, gender, and years of practice post MBBS as covariates for adjustment

Subgroup analyses will be performed based on experience with LLMs, gender and years of practice post MBBS. Scores for each case question will also be compared descriptively between the two randomized groups.

For a secondary exploratory analysis, cases completed solely by GPT-4o will be analyzed as a distinct third group using a nested design with three attempts per participant. Comparisons between GPT-4o cases and those from human participants will be conducted using mixed-effects modeling, with each case treated as an individual attempt under a participant using an analogous nested framework. While the study design minimizes the potential for missing data, if missing data does occur, multiple imputation will be considered under the intention-to-treat framework for the primary analysis. No interim analyses are planned given the small sample size.

All statistical analyses will be performed using Python software, version 3.11.12 (Python Software Foundation) with pandas for data manipulation and statsmodels version 0.14.4 for mixed-effects modeling. Statistical significance will be based on a p value.

***References***

1. Goh E, Gallo R, Hom J, et al. Large Language Model Influence on Diagnostic Reasoning: A Randomized Clinical Trial. *JAMA Netw Open.* 2024;7(10):e2440969. doi:10.1001/jamanetworkopen.2024.40969
